# Quantifying the social distancing privilege gap: a longitudinal study of smartphone movement

**DOI:** 10.1101/2020.05.03.20084624

**Authors:** Nabarun Dasgupta, Michele Jonsson Funk, Allison Lazard, Benjamin Eugene White, Stephen W. Marshall

**Affiliations:** University of North Carolina at Chapel Hill, 521 S. Greensboro Street, Carrboro, NC 27510 USA

## Abstract

**Background:** In response to the coronavirus pandemic, social distancing became a widely deployed countermeasure in March 2020. We examined whether healthier and wealthier places more successfully implemented social distancing.

**Methods:** Mobile device location data were used to quantify declines in movement by county (n=2,633) in the United States of America, comparing April 15–17 (n=65,544,268 traces) to baseline of February 17 - March 7. Negative binomial regression was used to estimate gradients of privilege across eleven healthcare and economic indicators, adjusting for rurality and stay-at-home mandates. External validation used separate venue-specific data from Google Location Services.

**Findings:** Counties without stay-at-home orders showed a mobility decline of −52·3% (95% CI: −50·3%, −54·3%), slightly less than the decline in mandated areas (−60·8%; 95% CI: −60·0%, −61·6%). Strong linear gradients in privilege were observed. After adjusting for rurality and stay-at-home orders, counties in the highest quintile of social distancing mobility restriction had: 52% less uninsured, 47% more primary care providers, 29% more exercise space, 27% less food insecurity, 26% less child poverty, 17% higher incomes, 14% less overcrowding, 9·6% more racial segregation, 8·2% less youth, 7·4% more elderly, and 6·2% less influenza vaccination, compared to least social distancing areas.

**Interpretation:** Healthier and wealthier counties displayed a social distancing privilege gap, measured via smartphone mobility change. Structural inequities in this key countermeasure will influence immunity, and disease incidence and mortality.

**Funding:** None

## Research in context

### Evidence before this study

We searched PubMed, Google Scholar, and a metasearch for pre-print servers search.bioPreprint, without language or date limits: (*coronavirus* OR *covid* OR *sars* OR *pandemic*) AND (*gps* OR *smartphone* OR *location services* OR *mobility*). Nine articles were found using mobility data to track changes during the COVID-19 pandemic, including analyses of the United States, Italy, and China. The only peer-reviewed article used a proprietary intercity travel index as model input for recreating the spread of SARS-CoV-2. No analysis of health disparities was evident. Additional searches for *social distancing* AND *privilege* returned zero results.

A Google News search (terms: covid AND mobility) identified eight primary data provider dashboards not reported in scientific literature, and dozens of derivative data visualizations. Only one data provider had a relevant metric, a “social inequality trend” defined as the mobility differential between wealthier and poorer neighborhoods within each metropolitan area.

### Added value of this study

This is the first large-scale study providing empirical evidence of financial and structural privilege on social distancing, using more than 65 million mobile device traces. We extend digital epidemiology methods by linking multiple data sources to adjust for potential confounding. We also conducted external validation of location data.

### Implications of all the available evidence

This study identifies specific systems of privilege that enable the ability to social distance. Success at social distancing during the pandemic was enabled by structural and economic factors that existed before pandemic start, consistent with anecdotal observations. Awareness of systems of privilege should inform the next phase of interventions and allocation of resources in order to ensure equity.

## INTRODUCTION

Concerns have been raised that compliance with stay-at-home orders (“social distancing”) might be a manifestation of social and economic privilege.^1,2^ However, there is limited systematic documentation of impacts of privilege during the coronavirus pandemic. This study examined whether gradients in structural and economic factors play a role in the adoption of social distancing, with inequities based on wealth, health, and power. We hypothesized that counties that were healthier or wealthier would adopt social distancing countermeasures with more intensity. Our hypothesis is based on the “healthy user effect,” which can be described as the propensity for patients who partake in one preventive measure to seek additional preventive services or engage in other healthy behaviors.^3,4^ We sought to quantify the social distancing privilege gap in order to suggest priorities for action.

*Social distancing* is physical separation to prevent disease transmission. In the United States of America (USA) and elsewhere, this has been implemented in the form of mass self-sequestration at home, accompanied by suspension of access to places of employment, education, recreation, and worship. In the absence of population-based testing or vaccination, social distancing has become the primary intervention for slowing viral transmission.

Aggregate movement data from mobile phones have emerged as a potential tool for understanding viral transmission, evaluating interventions, and monitoring compliance with countermeasures.^5^ These data could also be used to assess if pandemic response is poised to exacerbate underlying inequities in health. Location tracking includes global positioning system (GPS) data generated from mobile devices (smartphones, tablets, wearable activity trackers, in-vehicle navigation systems, etc.). In general, mobility traces can be generated using a device’s location via tracking apps or when switching signals are generated by mobile phone users moving between cell tower coverage areas. In the context of COVID-19, mobility data have been used as a proxy to measure intervention effects and model transmission patterns.^6–15^ In previous emergency responses they have been used to evaluate evacuation behavior after earthquakes^16^ and displacement after wildfires and cyclones.^17^ Legitimate concerns about privacy have also been raised.^18^

A recent report by Thompson and Serkez^19^ analyzed county-level smartphone mobility data to establish temporal trends in social distancing, stratifying by political leanings. However, this and other analyses did not control for rurality, which could be a strong explanatory factor in movement data. Therefore, we sought to identify healthcare and socioeconomic characteristics of places where social distancing was most successfully implemented within levels of rurality. Such analyses have the potential to inform messaging about social distancing. They also factor into the design of future studies evaluating the countermeasures.

## METHODS

This study linked established^20^ community-level health, social, and economic indicators with mobility data.^7^ Validation was conducted using a second mobility dataset with more granular information on venue.

### Baseline Health Data

In order to identify explanatory health and socioeconomic indicators, we used the 2019 Robert Wood Johnson Foundation County Health Rankings (CHR) dataset.^20,21^ The publicly available dataset contains dozens of metrics compiled from national surveys and healthcare databases.

We compared the variation in intensity of social distancing to eleven county-level metrics: three healthcare, two economic, three structural, and three demographic. These were selected from the CHR dataset because they are established indicators,^20^ with an emphasis on specific concerns arising during the pandemic.

### Healthcare metrics

To gauge overall baseline healthcare access, we examined primary care providers per 100,000 population and percent uninsured under age 65 (e.g., Medicare eligibility). We examined health insurance coverage among adults under age 65. As a marker for a related preventive health behavior, we also separately evaluated whether baseline influenza vaccination rates were associated with how much the county was likely to slow down during the current coronavirus outbreak, quantified as the percent of annual Medicare enrollees vaccinated.

### Economic metrics

We explored two baseline economic metrics, one representing overall community wealth (80^th^ percentile of annual household income in dollars) and one as proxy^22^ for child poverty (percent of school-age children eligible for subsidized or free lunches).^23^

### Structural metrics

Three lifestyle metrics were selected to provide a diverse range of baseline structural factors that could influence compliance with prolonged stay-at-home orders. The percent of people experiencing food insecurity was established from survey responses and a cost-of-food index. Opportunities for physical activity was modeled as the percent of population with adequate access to nearby parks and recreational facilities. The percent of households with overcrowding was based on housing condition surveys.

### Demographic metrics

The three demographic metrics were: percent of youth (age under 18 years) because of concerns about non-compliance with stay-at-home orders, the percent of elderly (aged 65 years and above) because they are a risk group for COVID-19 mortality, and a residential segregation index (white versus non-white) to address broader concerns about privilege and identity.^24^

### Primary Mobility Data

The analytic dataset started with public, aggregated county-level (or similar geopolitical unit assigned by coordinates of origin) data from smartphone GPS movement tracing, pre-processed by Descartes Labs (Santa Fe, New Mexico, USA). Raw mobility data generated from location services were processed using a parallel bucket sort to create device-based (e.g., node) records that for a given day were longitudinal.^25^ Maximum distance mobility (M_max_) was defined as the maximum Haversine (great circle) distance in kilometers from the first location report.^7^ Conceptually, this represents the straight-line distance between the first observation and the day’s farthest. Across all reports, the median accuracy of location measurement was 15 to 20 meters. Adjustments were made for poor GPS signal locks, too few observations (less than 10 reports per day), and signal accuracy (50 meter threshold).^7^ To reduce bias from devices merely transiting through (e.g., interstate highways), M_max_ is limited to nodes with at least 8 hours of observation per day. The resulting analysis dataset contained 2,633 of 3,142 US counties.

The pre-processed mobility dataset in this analysis could not be used to identify individuals. Using geotemporally coarse data provides further safeguards for protecting privacy. To this end, the values of M_max_ were summarized by taking the median per county-day (*m50*). The median was indexed against a baseline period to obtain our main study metric: the percentage change in mobility since baseline (*m50_index*) as a proxy for the intensity of social distancing. The baseline period of February 17 through March 7, 2020 was established by the data provider as a stable period before widespread social distancing.

### Variable Construction

To account for weekly periodicity in movement (e.g., less on weekends) we limited analysis to weekdays. To prevent undue influence from single-day variability and to allow two weeks to have elapsed since the last round of state stay-at-home orders, we averaged values of *m50_index* for the three most recent available weekdays at the time of analysis: April 15, 16, 17. The resulting distribution approximated a Gaussian function. To reduce outlier influence, we constructed a five-level inverted stratification of *m50_index*, where the highest quintile (5) represented the greatest reduction in mobility since baseline, interpreted as the highest 20% of counties in terms of social distancing intensity. Category boundaries for *m50_index* by quintile were: lowest mobility change (1) +193% to −45·8%, (2) −46·0% to −55·4%, (3) −55·5% to −62·3%, (4) 62·4% to 74·8%, and highest (5) −75·0% to −100%. Only seven counties (all less than 20,000 population) showed an *increase* in mobility from baseline and were included in the lowest quintile.

### Potential Confounders

We adjusted for two potential confounders. First, state and municipal stay-at-home-orders from February through April 2020 were classified by county.^26^ These orders limited travel to basic necessities and employment in sectors deemed essential. Municipal stay-at-home orders were accounted for in eight states without mandates.

Second, even though our main outcome was *change* in mobility from baseline, rurality might be a potential confounder due to distances traveled for essential activities. We used federal rural-urban continuum codes (RUCC) to adjust for rurality and transportation connections between city centers and satellite counties.^27^ Although RUCC can be conceptualized as a 9-point ordinal scale of urbanicity (or rurality), it was modeled using indicator coding to impose fewer assumptions. Other potential spatial and economic confounders (number of solo vehicle commuters, long commuting times, and composite socioeconomic indices) were not included because they did not meaningfully improve model fit.

### External Validation

In the context of social distancing, general movement data have the potential for misclassification. One way to validate the findings is to compare these data to more granular location information, such as by type of visited venue. The data came from aggregated and anonymized GPS traces of devices for which the Location History setting within Google apps had been turned on enabled. Since only limited information on data collection was available,^11^ we did not consider the Google Location Services data appropriate for the primary analysis at this time.

During the study period, Google LLC (Mountain View, California, USA) published county-level datasets^28^ showing COVID-19-related mobility changes across six types of venues: grocery and pharmacy; parks, transit stations, retail and recreation, places of residence; and places of work. The metric was percent change in mobility since baseline, January 3 to February 6, 2020, controlling for day of week. A validation dataset was created for March 1 to April 11, the overlap period with mobility data used in the primary analyses, for which county-day could be established. To have confidence in overall mobility change to serve a proxy for social distancing, we expected the strongest correlations with staying at home, transit, work and retail, and less correlation with other venues that were permissible or essential.

### Statistical Analysis

Datasets were analyzed with Stata MP (version 16, College Station, Texas, USA). Scaled Poisson regression with robust variance estimators was used in base models regressing each of the 11 metrics individually against quintiles of mobility change. Negative binomial (NB2) models were employed when warranted by residual overdispersion. The adjusted models included indicator variables to control for rurality/urbanicity and governmental stay-at-home mandates. For a given health or socioeconomic indicator, mean and 95% confidence intervals were calculated for each quintile using non-intercept models; pairwise contrasts of percent difference between quintiles were estimated using the full model with adjustment. For validation we compared zero-recentered *m50_index* against percent change from baseline by venue, as reported by Google Location Services users. Structural equation modeling was used to calculate pairwise correlations and 95% confidence intervals, accounting for repeated measures. Code and datasets are available at https://github.com/opioiddatalab/covid.

### Ethics Statement

The aggregated public county-level data analyzed in this study were not considered human subjects research and exempted from ethics review per the guidance of the University of North Carolina Office of Human Research Ethics.

## RESULTS

We analyzed data from 65,544,268 mobile GPS traces for the 3-days April 15, 16, 17 of 2020, comparing mobility to an early COVID-19 baseline period (February 17 through March 7). These traces came from 2,633 US counties and each device was observed for more than 8 hours.

### Mobility Changes

Across the United States, there was an average decline of −59·7% in mobility from baseline. Mobility decline ranged from −86·5% in the quintile with the most restricted movement, and −33·2% in the least (Table 1). The 337 counties *without* stay-at-home orders showed a decline in movement of −52·3% (95% CI: −50·3%, −54.3%), while the 2,296 counties where stay-at-home orders had been enacted experienced a slightly greater decline, −60·8% (95% CI: −60·0%, −61·6%).

**Table 1.**
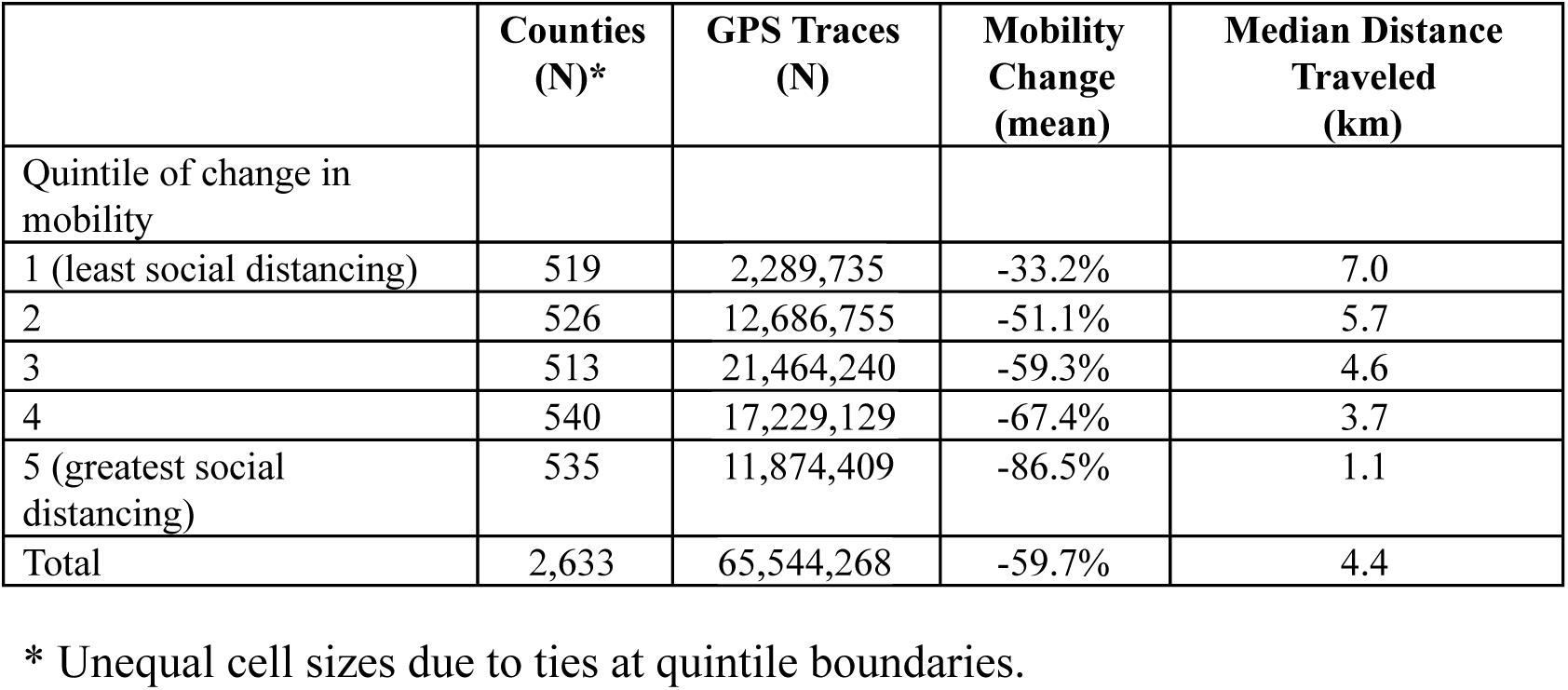
Average change in GPS-measured movement from baseline, by quintile of mobility change, April 15 to 17, 2020, United States.

In general, counties that reduced movement tended to be more urban, however, there were many exceptions to this trend. For example, 33% (n=137) of counties in the highest tier of social distancing had less than 20,000 population (RUCC 6 or greater), found in states with earlier (late March) stay-at-home orders like Michigan, Minnesota, and Texas. Similarly, 107 (25%) metropolitan counties (RUCC 3 or lower) were found in places with less mobility curtailment, for example in states with later (early April) stay-at-home orders most commonly Georgia, Louisiana, and Alabama. All subsequent results in this paper are adjusted for rurality/urbanicity and the enactment of stay-at-home orders.

### Healthcare Indicators

All eleven explanatory healthcare, economic, structural and demographic factors consistently showed near-linear associations with intensity of social distancing as measured through mobility declines, Figure 1. The counties with the smallest declines in mobility had 50 primary care providers per 100,000, whereas the most social distancing counties had 74 per 100,000 after adjusting for rurality and stay-at-home orders, a 47% (95% CI: 38%, 58%) difference. Counties with lower social distancing had a higher proportion of people without health insurance. The lowest social distancing counties had 10·7% uninsured, whereas the most social distancing counties had only 7·0% uninsured, a 52% (95% CI: 45%, 60%) difference. Overall county influenza vaccination rates among Medicare beneficiaries varied from 9% to 65% at baseline, with a mean of 42%. On average, counties with more social distancing had 6·2% (95% CI: 3·7%, 8·7%) higher flu vaccinations at baseline.

**Fig 1.**
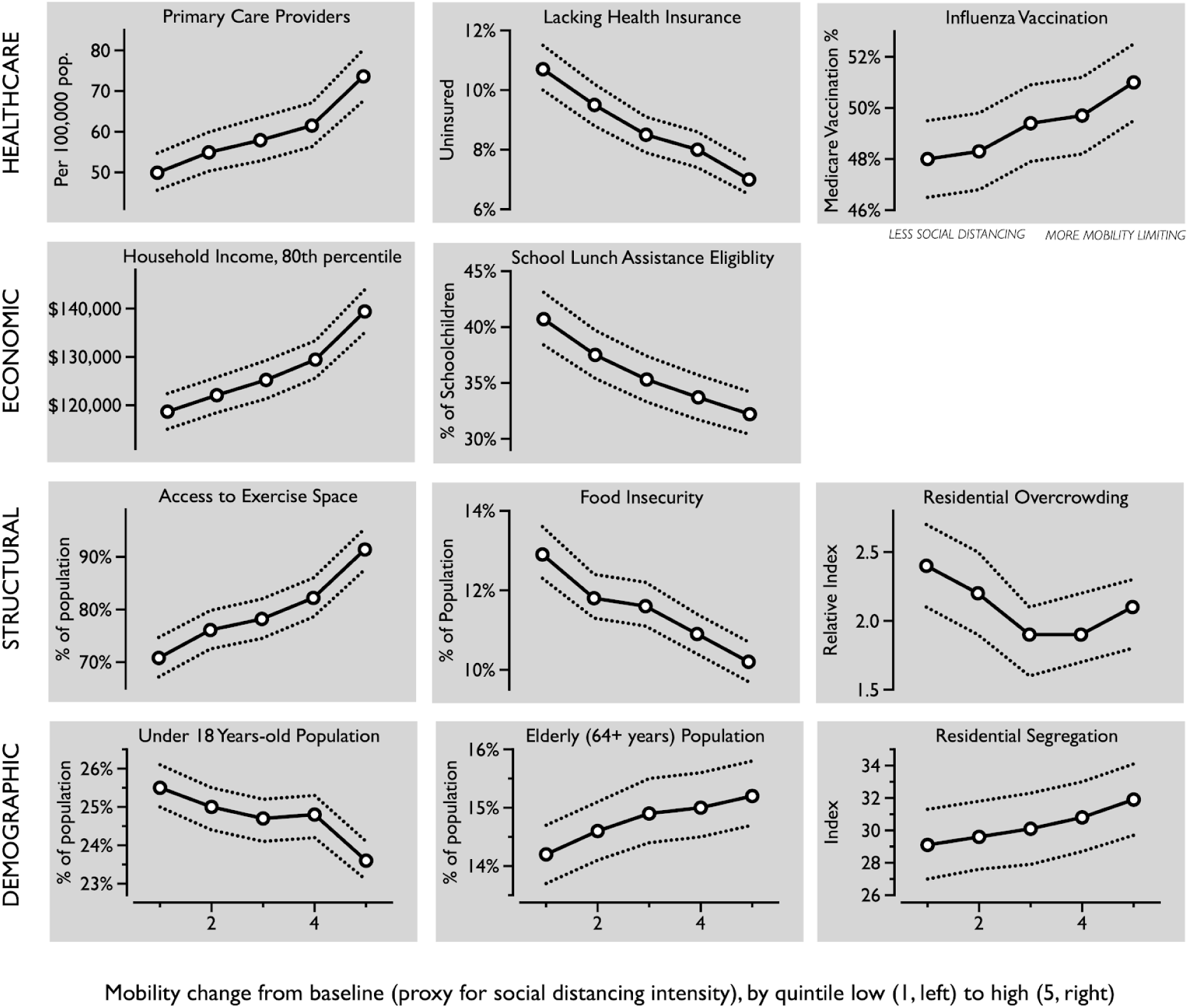
US Counties (N=2,633) with greater privilege in healthcare resources and wealth adopted social distancing with greater intensity, as measured by reductions in mobile device movement. Horizontal axis increases in social distancing intensity by quintile from left (1) to right (5), adjusted for rurality and stay-at-home orders.

### Economic Indicators

There was a strong increasing trend between income and intensity of social distancing. Places that were able to most curtail movement had 17% (95% CI: 15%, 20%) higher annual household income, after adjusting for rurality and social distancing orders. In places with the least mobility decline, the 80th percentile of annual household income was around $120,000, whereas in the most social distancing counties it was $140,000. Correspondingly, 41% of school age children were eligible for free or subsidized lunches in the least social distancing areas, which was 26% (95% CI: 21%, 31%) higher than in areas that were more able to restrict movement.

### Structural Indicators

The lowest social distancing counties had greater food insecurity, among 13% of residents. The most social distancing counties had 10%. After adjusting for rurality and social distancing orders, this amounted to a 27% (95% CI: 23%, 32%) difference. In the lowest social distancing counties, only 71% of residents had access to physical spaces for exercise, whereas in the most social distancing counties 91% had access, a 29% (95% CI: 24%, 34%) difference. The most social distancing counties had 14% (95% CI: 4·4%, 25%) less overcrowding, but showed greater variability.

### Demographic Indicators

Counties with the least social distancing were younger: 8·2% more youth (95% CI: 6·4%, 10%) younger than 18 years. Counties that did the best at social distancing were older, with 7·4% (95% CI: 4·7%, 10%) more elderly people compared to the lowest tier. Counties that limited mobility the most also had 9·6% higher (95% CI: 4·1%, 15%) residential racial segregation (white compared to non-white).

### External Validation

County-level mobility and venue-specific changes were well-correlated, Table 2. Retail and recreation, work, and transit showed positive linear correlations above 0·70. Changes in the use of parks (0·24) and grocery/pharmacy (0·53) were less correlated with overall mobility. Conversely, and as anticipated, changes in mobility showed a strong (negative) correlation with increases in staying at home (−0·83). These results lend further credence that using overall mobility change as a proxy for social distancing was justified.

**Table 2.** Correlation between changes in overall mobility versus venue-specific location information from Google Location Services, March 1 through April 11, 2020.

|  | County-day Frequency* | Correlation (95% CI) |
| --- | --- | --- |
| **Retail and recreation**<br>restaurants, cafes, shopping centers, theme parks, museums, libraries, movie theaters | 92,681 | 0.765 (0.762, 0.767) |
| **Grocery and pharmacy**<br>grocery markets, food warehouses, farmers markets, specialty food shops, drug stores, pharmacies | 89,059 | 0.535 (0.530, 0.540) |
| **Places of work** | 101,290 | 0.747 (0.744, 0.750) |
| **Homes and residences** | 49,666 | -0.830 (-0.832, -0.827) |
| **Parks**<br>local parks, national parks, public beaches, marinas, dog parks, plazas, public gardens | 27,182 | 0.242 (0.231, 0.254) |
| **Transit**<br>public transportation hubs for subway, bus, and trains | 41,299 | 0.709 (0.704, 0.713) |

\* Varying cell sizes due to low count suppression in the Google Location Services dataset.

## DISCUSSION

Counties that were most successful at limiting mobility through social distancing were healthier and wealthier before pandemic onset. This paper provides empirical evidence that places with greater social distancing had distinctive healthcare, economic, and structural advantages that may have facilitated compliance with stay-at-home orders. Awareness about barriers to social distancing faced by low wage workers has been recognized.^28^ However, much less awareness exists about privilege’s consequences. Consistent with the concept of privilege as a system, our analysis documents the specific advantages of privilege in a crisis situation.^29^

Of the eleven metrics we examined, one of the starkest differences between the highest and lowest social distancing counties was in primary care providers per capita, 47%. But, influenza vaccination among Medicare beneficiaries showed only a 6% difference. Over decades, a vaccine initially intended for primary care settings was expanded to allow administration by nurses and pharmacists, benefitting from streamlined record keeping and reimbursement processes, no additional patient cost (in Medicare), and administration venues outside of traditional healthcare delivery. If a coronavirus vaccine becomes available, one way to close the social distancing privilege gap could be to make it at least as easy to obtain as the flu vaccine.

Since the flu vaccine is free to all Medicare beneficiaries, and the elderly group has the most influenza mortality, this might represent how health conscious the population was, on average. However, in the Medicare population not getting a flu vaccine is also a marker for frailty. Failure to receive the flu vaccine, even when there is no influenza circulating, is a strong predictor of mortality.^29^ An alternative interpretation is that places with low social distancing could have a greater share of the elderly who are exceptionally vulnerable.

A key concern during pandemic quarantine is how to maintain routine healthcare screening for chronic diseases. In-person clinic visits have been curtailed drastically in favor of telemedicine, but the replacement is not complete. Blood pressure cannot be readily checked remotely. While misclassification could arise if patients routinely cross county lines to see doctors (and providers may cluster together within health systems), our observations suggest that longstanding inequities in chronic disease screening and testing may also be exacerbated. The 52% difference in health insurance coverage will also need to be addressed.

The observed 17% income differential cannot be explained by location alone, since we controlled for urbanicity. Food insecurity ranked highly in disparity between high and low social distancing counties. As evidenced on social media, a phenomenon of the quarantine era has been for otherwise busy professionals to take up labor intensive food projects, epitomized by baking bread. At the same time, food banks have been expending stocks and asking for donations. Granular demographic information is routinely used to customize social media messages in advertising. Perhaps our findings can serve to inspire new avenues for tailored public health messaging to reduce the social distancing privilege gap.

As societies weigh options for removing restrictions on social distancing, our findings encourage considering ways to even social distancing ability. Epidemiologic models for betacoronavirus transmission suggest that social distancing measures may need to be selectively loosened, and possibly reinstated, over the next two years.^30^ Assuming seasonal forcing in the autumn, these models make clear that in the absence of pharmaceutical intervention, the surest way to manage hospital load is through controlled cultivation of population immunity. We found that voluntary sequestration rates were similar to areas with stay-at-home mandates, and that racial segregation was highest in areas with most social distancing. This suggests that those wealthy enough to remain cocooned may minimize their exposure to the virus even after stay-at-home orders are lifted. In such a scenario, the bulwark of herd immunity would likely be achieved through infection of less privileged members of society. While durability of immunity and asymptomatic carriage of SARS-CoV-2 are unknown, the circumstances are set for those with the greatest privilege to benefit from communal protection, while incurring a lower burden of disease.

### Limitations

We measured *change in movement* as a proxy for success in implementing social distancing. A limitation is that change in movement does not capture physical proximity to other people. Maintaining two meters of distance from others is a key element of social distancing not reflected in mobility changes measured in kilometers. Our study also has an underrepresentation of extremely rural areas because we required at least ten 8-hour smartphone traces per day to maintain privacy; findings might not be generalizable to these areas. The distributions of jobs that are amenable to remote work-from-home likely also vary by rurality, but we did not have data to test this hypothesis beyond commute times. We intentionally did not analyze positive-among-tested coronavirus counts because of strong variation between states leading to selection bias (e.g., diagnostic suspicion bias for test eligibility).

## CONCLUSIONS

This study is innovative in using smartphone location tracking for measuring the extent of public health interventions on a national scale. We found that the adoption of social distancing practices was strongly correlated with better community health and financial resources. Our findings also suggest the need to innovate additional measures to prevent viral transmission in places where social distancing is less feasible. Awareness of systems of privilege should inform the next phase of pandemic interventions and provision of healthcare resources in order to ensure equity.

## Data Availability

The study used only data that were publicly available.

https://github.com/opioiddatalab/covid

## Acknowledgments

We thank David Joerg for providing access to formatted data. We thank Anindita Dasgupta and Roxanne Saucier for helpful editorial comments.

## Author Contributions

BW processed source data. ND conducted the analysis and drafted the first version of the manuscript, and is the party responsible for data integrity. MJF provided the impetus for the initial study and provided clinical interpretations during analysis. AL and SWM contributed to the drafting of the manuscript and interpretation of results.

## Data and materials availability

All code and data available at https://github.com/opioiddatalab/covid.

